# Reach and effectiveness of a HEARTS hypertension control pilot project in Guatemala

**DOI:** 10.1101/2024.04.03.24305304

**Authors:** Irmgardt Alicia Wellmann, José Javier Rodríguez, Benilda Batzin, Guillermo Hegel, Luis Fernando Ayala, Kim Ozano, Meredith P. Fort, Walter Flores, Lesly Ramirez, Eduardo Palacios, Mayron Martínez, Manuel Ramirez-Zea, David Flood

## Abstract

The World Health Organization’s HEARTS Technical Package aims to improve the primary care management of hypertension and other cardiovascular risk disease factors at the population level. This study describes the first HEARTS implementation project in the Ministry of Health primary care system in Guatemala. This pilot project was implemented from April to December 2022 in 6 primary health facilities in 3 rural, Indigenous municipalities. The project consisted of HEARTS-aligned strategies that were adapted to foster program sustainability in Guatemala. Outcomes were defined using the RE-AIM framework. The primary *reach* outcome was treatment rate, defined as the absolute number of patients each month receiving medication treatment for hypertension. The primary *effectiveness* outcomes were mean systolic blood pressure (BP), mean diastolic BP, and proportion with BP control. In the first month of the post-implementation period, there was a significant increase of 25 patients treated (P=0.002), followed by a significant increase thereafter of 2.4 additional patients treated each month (P=0.007). The mean change in systolic BP was -4.4 (95 CI -8.2 to -0.5, P=0.028) mmHg and mean change in diastolic BP was -0.9 (95 CI -2.8 to 1.1, P=0.376) mmHg. The proportion of the cohort with BP control increased from 33.4% at baseline to 47.1% at 6 months (adjusted change of 13.7% [95 CI 2.2% to 25.2%, P=0.027]). These findings support the feasibility of implementing the HEARTS model for blood pressure control in the Guatemalan MOH primary care system where the vast majority of hypertension patients seek care.

## Introduction

The World Health Organization’s HEARTS Technical Package aims to improve the primary care management of hypertension and other cardiovascular risk disease (CVD) factors at the population level.^1^ The Pan American Health Organization has spearheaded efforts to implement HEARTS in health systems in 33 countries in the Americas region (20 belong to Latin America and 13 to the Caribbean) through its “Hearts in the Americas” initiative starting in 2016 with the first cohort.^2^ This study describes the first HEARTS implementation project in Guatemala. Guatemala is a lower middle-income country and the most populous nation in Central America. Approximately 80% of the population in Guatemala is uninsured and dependent on the Ministry of Health (MOH) for primary health care.^3^

## Methods

The HEARTS pilot in Guatemala was implemented from April to December 2022 in 6 primary health facilities in 3 rural, Indigenous municipalities in the department of Sololá. The combined population in the municipalities is approximately 15,500 people. The MOH was the implementing institution, and the Institute of Central America and Panama (INCAP) provided technical assistance and evaluated the project. The Center for the Study of Equity and Governance in Health Systems (CEGSS) provided technical assistance for the establishment and training of self-help group of patients. Funding support was provided by Resolve to Save Lives. Ethics approval was obtained from the MOH and INCAP.

The implementation project consisted of the following HEARTS-aligned strategies adapted to Guatemala,^4^ as well as patient-centered strategies to foster program sustainability: (1) Coordination with MOH leadership to ensure availability of antihypertensive medications and blood pressure (BP) monitoring devices; (2) dissemination and training on standardized hypertension treatment protocols; (3) task sharing, as nurses (professional and auxiliary) primarily delivered care under physician supervision; (4) implementation of an electronic monitoring tool, the District Health Information System 2 (DHIS2) system; (5) establishing and training of community self-help groups to generate patient demand for MOH services and to monitor the MOH’s implementation; and (6) implementation of a municipal pharmacy with low-cost and generic antihypertensive medications.

We used the RE-AIM framework to guide our implementation evaluation of this HEARTS pilot project.^5^ This letter reports reach and effectiveness outcomes. Other RE-AIM outcomes (adoption, implementation, and maintenance) will be reported in future studies.

The primary *reach* outcome was treatment rate, defined as the absolute number of patients each month receiving medication treatment for hypertension. Patients who were pregnant or were treated for acute hypertension were excluded from this calculation. The MOH requires medications to be refilled monthly, so this outcome is a meaningful indicator of population coverage. The data source for this outcome was the MOH’s Health Management Information System (*Sistema de Información Gerencial de Salud* [SIGSA]), which tracks dispensed medications with very high fidelity. We analyzed these data using a single-group interrupted time series approach with segmented linear regression and Newey-West standard errors to account for autocorrelation.

The primary *effectiveness* outcomes were mean systolic BP, mean diastolic BP, and proportion with BP control (<130/80 mmHg, aligning with local guidelines^6^). The data source for this outcome was a panel of 102 patients with hypertension recruited at baseline and monitored over 6 months. Potentially eligible patients were recruited from MOH records, referrals from community members, and screening through home visits. Eligibility criteria were age ≥20 years and either (a) previously diagnosed hypertension; (b) BP ≥140/90 mmHg; or (c) BP ≥130/80 if also taking an antihypertensive medication, having a 5-year CVD risk ≥10%,^7^ or reporting a history of CVD. We used a blood pressure measurement protocol reported previously.^4^ We analyzed these data using a pre-post approach with multilevel linear and logistic regression models for continuous and dichotomous outcomes, respectively. We specified a random intercept for participant and fixed effects for intervention time, municipality, age, and sex. Analyses were done in Stata version 17.

## Results

### Reach

The Figure shows monthly treatment rate in the health district in the 15 months before the HEARTS pilot and the 16 months following implementation. In the pre-implementation period, approximately 20-25 hypertension patients were treated per month with no significant monthly trend. In the first month of the post-implementation period, there was a significant increase of 25 patients treated (P=0.002), followed by a significant increase thereafter of 2.4 additional patients treated each month (P=0.007). By month 16 post-implementation, approximately 80 hypertension patients were treated per month.

**Figure:**
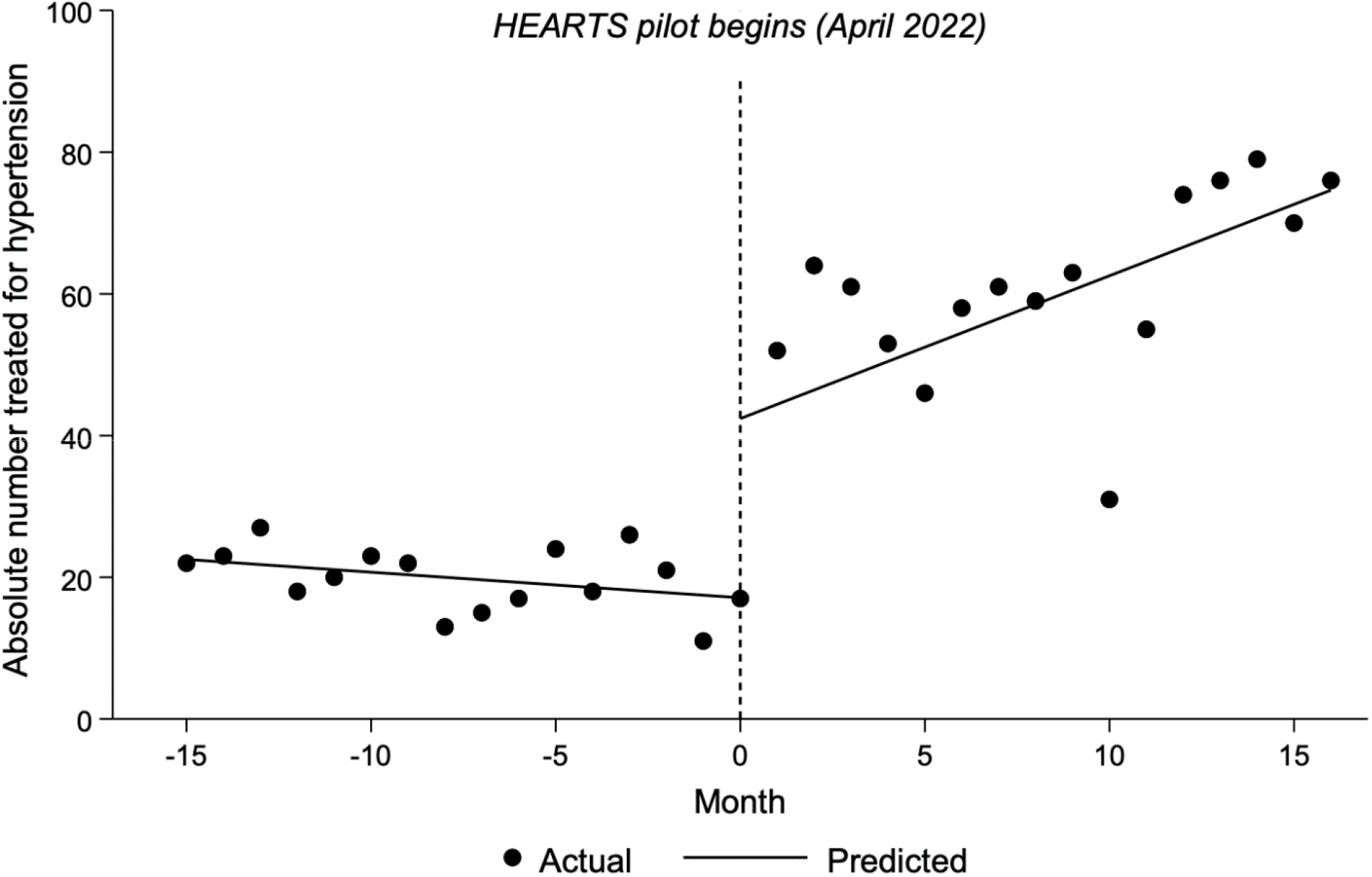
Reach outcomes of monthly treatment rate. Note: Data underlying this figure were obtained from the Guatemala MOH’s Health Management Information System. Lines reflect the single-group interrupted time series approach with segmented linear regression.

### Effectiveness

In the hypertension panel, 85% of participants were women and median age was 67 years (interquartile range 56 to 75 years). The Table shows blood pressure results over 6 months. The mean change in systolic BP was -4.4 (95 CI -8.2 to -0.5, P=0.028) mmHg and mean change in diastolic BP was -0.9 (95 CI -2.8 to 1.1, P=0.376) mmHg. The proportion of the cohort with BP control increased from 33.4% at baseline to 47.1% at 6 months (adjusted change of 13.7% [95 CI 2.2% to 25.2%, P=0.027]).

**Table:** Effectiveness outcomes of mean blood pressure and % blood pressure control in n=102 participants.

| **Outcome** | **Baseline**<br>**(95% CI)** | **6 months**<br>**(95% CI)** | **Adjusted**<br>**difference**<br>**(95% CI)** | **P value** |
| --- | --- | --- | --- | --- |
| Systolic blood pressure, mmHg | 134.8<br>(131.1 to 138.4) | 130.4<br>(126.8 to 134.1) | -4.4<br>(-8.2 to -0.5) | 0.028 |
| Diastolic blood pressure, mmHg | 72.1<br>(70.1 to 74.1) | 71.2<br>(69.3 to 73.2) | -0.9<br>(-2.8 to 1.1) | 0.376 |
| Blood pressure control of <130/80, % | 33.4<br>(24.6 to 43.1) | 47.1<br>(37.9 to 56.4) | 13.7<br>(2.2 to 25.2) | 0.027 |

Note: Data underlying this table were obtained from a panel of 102 patients with hypertension recruited at baseline and monitored over 6 months. Data were analyzed using multilevel linear and logistic regression models for continuous and dichotomous outcomes, respectively

## Discussion

In this small-scale HEARTS implementation pilot project in the MOH in Guatemala, we observed a 4-fold increase in the treatment rate and a 14% increase in the proportion achieving BP control. These findings support the feasibility of implementing the HEARTS model for blood pressure control in the Guatemalan MOH primary care system where the vast majority of hypertension patients seek care.

An important secondary finding was the extremely low baseline coverage of MOH hypertension treatment, with just 20 patients in a catchment area with an estimated total number of 1,500 people with hypertension (approximately 1% population treatment coverage at baseline). While the 4-fold increase in treatment observed in this pilot is noteworthy, there is still a critical need to dramatically expand coverage at the population level. In addition to efforts to improve medication availability, we are pursuing other strategies such as standardizing 90-day prescription refills in the MOH, as recommended by HEARTS experts.^8^

Since our pilot was launched in April 2022, the Guatemalan MOH has officially pledged to implement HEARTS.^9^ We are now focusing on supporting the MOH in its implementation efforts across the country. We also are developing strategies to integrate the primary care management of other CVD risk factors such as diabetes into the HEARTS hypertension model.^10^ Finally, we hope to generate evidence to support HEARTS implementation projects in other countries, including making causal estimates of impact, providing guidance on adapting HEARTS to new health systems, and calculating cost-effectiveness.

## Data Availability

All data produced in the present study are available upon reasonable request to the authors

## References

1. WHO. Hearts: Technical package for cardiovascular disease management in primary health care. Geneva: World Health Organization, 2016.

2. PAHO. HEARTS in the Americas. 2024. https://www.paho.org/en/hearts-americas (accessed February 27, 2024).

3. Instituto Guatemalteco de Seguridad Social. Informe anual de labores 2023. Guatemala; 2023.

4. Paniagua-Avila A, Fort MP, Glasgow RE, et al. Evaluating a multicomponent program to improve hypertension control in Guatemala: study protocol for an effectiveness-implementation cluster randomized trial. Trials 2020; 21(1): 509.

5. Glasgow RE, Harden SM, Gaglio B, Rabin B, Smith ML, Porter GC, et al. RE-AIM Planning and Evaluation Framework: Adapting to New Science and Practice With a 20-Year Review. Front Public Health. 2019 Mar 29;7. https://www.ncbi.nlm.nih.gov/pmc/articles/PMC6450067/ (accessed March 27, 2024).

6. Ministerio de Salud Pública Asistencia Social. Normas de Atención Salud Integral Para Primero y Segundo Nivel 2018: MSPAS, 2018.

7. Gaziano TA, Young CR, Fitzmaurice G, Atwood S, Gaziano JM. Laboratory-based versus non-laboratory-based method for assessment of cardiovascular disease risk: the NHANES I Follow-up Study cohort. The Lancet 2008; 371(9616): 923–31.

8. Brettler JW, Arcila GPG, Aumala T, et al. Drivers and scorecards to improve hypertension control in primary care practice: Recommendations from the HEARTS in the Americas Innovation Group. The Lancet Regional Health - Americas 2022; 9.

9. Pan American Health Organization. Implementarán iniciativa Hearts para la prevención y el control de las enfermedades cardiovasculares (ECV) en Guatemala. 2022. https://www.paho.org/es/noticias/11-11-2022-implementaran-iniciativa-hearts-para-prevencion-control-enfermedades (accessed February 27, 2024).

10. Wellmann IA, Ayala LF, Rodríguez JJ, et al. Implementing integrated hypertension and diabetes management using the World Health Organization’s HEARTS model: protocol for a pilot study in the Guatemalan national primary care system. Implementation Science Communications 2024; 5(1).

